# Hospital readmissions of discharged patients with COVID-19

**DOI:** 10.1101/2020.05.31.20118455

**Authors:** Lina Marcela Parra Ramírez, Mireia Cantero Caballero, Ignacio Morrás de la Torre, Alberto Vallejo Plaza, Itziar Diego Yagüe, Elena Jimenez Tejero, Elena Múñez Rubio, Ángel Asensio, Ana Fernández Cruz, Antonio Ramos Martínez

## Abstract

**Background:** COVID-19 infection has led to an overwhelming effort by health institutions to meet the high demand for hospital admissions.

**Aim:** To analyse the clinical variables associated with readmission of patients who had previously been discharged after admission for COVID-19.

**Design and methods:** We studied a retrospective cohort of patients with laboratory-confirmed SARS-CoV-2 infection who were admitted and subsequently discharged alive. We then conducted a nested case-control study paired (1:1 ratio) by age, sex and period of admission.

**Results:** Out of 1368 patients who were discharged during the study period, 61 patients (4.4%) were readmitted. Immunocompromised patients were at increased risk for readmission. There was also a trend towards a higher probability of readmission in hypertensive patients (p=0.07). Cases had had a shorter hospital stay and a higher prevalence of fever during the 48 hours prior to discharge. There were no significant differences in oxygen levels measured at admission and discharge by pulse oximetry intra-subject or between the groups. Neutrophil/lymphocyte ratio at hospital admission tended to be higher in cases than in controls (p=0.06). The motive for readmission in 10 patients (16.4%), was a thrombotic event in venous or arterial territory (p<0.001). Neither glucocorticoids nor anticoagulants prescribed at hospital discharge were associated with a lower readmission rate.

**Conclusions:** The rate of readmission after discharge from hospital for COVID-19 was low. Immunocompromised patients and those presenting with fever during the 48 hours prior to discharge are at greater risk of readmission to hospital.

## Introduction

The dramatic effects of the COVID-19 pandemic on the population have overwhelmed many health institutions around the world. During several weeks, the demand for hospital beds has exceeded the capacity to admit patients (1). In addition, the high hospital attendance rate has reduced the capability to treat other serious diseases such as neoplasms or cardiovascular diseases. (2,3). Therefore, making the most efficient use of hospital beds should be an imperative objective during outbreaks of this nature.

Clear guidelines on when to discharge patients admitted with this infection have not yet been established. Therefore, physicians may act too cautiously, protracting hospital stay unnecessarily, or conversely, they may be too bold in discharging patients quickly, potentially worsening their prognosis. Although the study of re-admitted patients with COVID-19 is a very relevant problem, to date very little attention has been devoted to its different clinical aspects (5–7).

Therefore, we planned a study to analyse the clinical variables associated with readmission of patients who had previously been discharged after admission for COVID-19.

## Methods

### Setting

The study was conducted in a 613-bed tertiary care university hospital in Madrid (Spain). The first case of SARS-CoV-2 infection was confirmed the February 25^th^, 2020 in the Community of Madrid. Since then, this region accounts for 68 696 confirmed cases, 42 928 inpatients and 14 671 deaths, being the most affected area in Spain. In our hospital, the first case was identified February 27^th^, there have been nearly 1 500 in-patients, and the highest count of admitted patients was 626 which was reached on March 30^th^.

### Study Population

We studied a retrospective cohort of patients with laboratory-confirmed SARS-CoV-2 infection who were admitted and subsequently discharged alive. The study period extended between 26^th^ February 2020 to 20^th^ April 2020. SARS-CoV-2 was detected by a real-time PCR assay targeting E-gene, RdRP-gene and N-gene, performed with the protocol reported by the WHO (8)

### Case-Control study

We conducted a nested case-control study matched (ratio 1:1) by age, sex and period of admission. Two periods were defined based on the incidence of admission cases. The first covered from 26^th^ February to 26^th^ March, the period of highest cumulative incidence of admission cases, and the second from 27^th^ March to 20^th^ April.

A case-patient was defined as a patient with confirmed SARS-CoV-2 infection who had been readmitted within three weeks of discharge and clinical presentation of readmission was related with the infection or its treatment. Patients who were discharged but not readmitted were considered as controls, after excluding death during that period.

The medical data included parameters such as obesity, cancer, diabetes, chronic obstructive pulmonary disease (COPD), immunosuppression and hypertension. Obesity was defined as a body mass index greater than 30 kg/m2. Immunocompromised patients included those with solid organ or hematopoietic stem cell transplantation, HIV infection, or who were previously treated with glucocorticoids (equivalent dose of prednisone ≥ 15 mg / day) or immunosuppressive drugs. Symptoms (including their length), physical examination findings and laboratory measurements were extracted from the electronic medical record. Analytical data (hemogram, basic biochemistry, lactated hydrogenase, C-reactive protein, D-dimer and ferritin) at admission and discharge was recorded. To evaluate the radiological evolution, we consider the corresponding radiologist report. Outcomes included death and hospital discharge

Clinical presentations related with the infection or complications related to treatment for SARS-CoV-2 infection were registered as reasons for admission, including respiratory manifestations, venous or arterial thrombosis, exacerbation of chronic disease, organ failure, bacterial superinfection.

### Statistical analysis

The chi-squared test and Fisher’s exact test were used to compare categorical variables, and continuous variables by Student’s t-test and the Mann-Whitney U Test. Univariate and multivariate logistic regression was undertaken with Stata 13.0 software (StataCorp, College Station, US). All tests of significance were two-tailed, and values of p<0.05 were considered statistically significant.

## Results

Out of 1368 patients who were discharged during the study period, 61 patients (4.4%) were readmitted (table 1). The median time from discharge to readmission was 6 days (IQR 3-10). One hundred and sixty-three patients (12.5%) died during the first admission.

**Table 1.** Clinical characteristics of all COVID-19 patients admitted during the study period.

| Characteristics | Patient with a single admission(n=1307) | Readmitted patients (n=61) | p |
| --- | --- | --- | --- |
| Age (median, IQR) | 64 (54-75) | 67 (59-76) | 0.14 |
| Masculine gender (n, %) | 827 (63.3) | 45 (73.8) | 0.10 |
| Length of symptoms (median, IQR) | 7 (4-10) | 6 (3-10) | 0.43 |
| Admission to ICU (n, %) | 93 (7.1) | 3 (4.9) | 0.51 |
| Non-survivors (n, %) | 163 (12.5) | 9 (14.7)<sup>1</sup> | 0.61 |

IQR: interquartile range. ICU: Intensive care unit. <sup>1</sup>Patients who died during hospital readmission.

### Clinical characteristics of the patients who are readmitted

Table 2 displays demographic and clinical characteristics of cases and controls. Immunocompromised patients were at increased risk for readmission. There was also a trend towards a higher probability of readmission in hypertensive patients (p=0.07). Cases had had a shorter hospital stay and a higher prevalence of fever during the 48 hours prior to discharge. There were no significant differences in oxygen levels measured at admission and discharge by pulse oximetry intra-subject or between the groups.

**Table 2.** Demographic and clinical characteristics in cases and controls.

| Characteristic | Cases (n=61) | Controls (n=61) | p |
| --- | --- | --- | --- |
| Age (median, IQR) | 67 (59-76) | 66 (57-76) | 0.68 |
| Male (n,%) | 45 (73.8) | 45 (73.8) | 1.00 |
| Comorbidity |  |  |  |
| Obesity | 6 (9.8) | 5 (8.2) | 0.75 |
| Diabetes mellitus | 14 (22.9) | 10 (16.4) | 0.36 |
| Hypertension | 34 (55.4) | 24 (39.3) | 0.07 |
| Cardiovascular disease | 16 (26.2) | 12 (19.7) | 0.39 |
| COPD | 12 (19.7) | 12 (19.7) | 1.00 |
| Neoplasia | 12 (19.7) | 12 (19.7) | 1.00 |
| Immunosuppression<sup>1</sup> | 10 (16.4) | 3 (4.9) | 0.04 |
| Admission |  |  |  |
| Length of symptoms (median, IQR) | 6 (3-10) | 7 (4-9) | 0.52 |
| Admission to ICU (n, %) | 3 (4.9) | 5 (8.2) | 0.72 |
| Total hospital stay (median, IQR) | 6 (4-14) | 9 (6-14) | 0.02 |
| Pneumonia on admission (n, %) | 53 (86.9) | 59 (96.7) | 0.05 |
| Oxygen saturation (% mean, SD) | 94.9 (2.7) | 94.7 (2.0) | 0.68 |
| Discharge |  |  |  |
| Fever 48 hours at discharge (n, %) | 11 (18.0) | 4 (6.6) | <0.001 |
| Afebrile at discharge (days, median, IQR) | 5 (3-10) | 7 (5-11) | 0.03 |
| Oxygen saturation (% mean, SD) | 93.7 (12.4) | 94.9 (2.2) | 0.46 |
| Radiological evolution (n, %) |  |  |  |
| No change | 21 (42.9) | 18 (38.3) | 0.31 |
| Worsening | 16 (32.7) | 11 (23.4) |  |
| Improving | 12 (24.4) | 18 (38.3) |  |
| Glucocorticoid treatment at discharge (n, %) | 30 (49.2) | 26 (42.62) | 0.47 |
| Anticoagulants at discharge<sup>2</sup> (n, %) | 16 (26.2) | 17 (28.9) | 0.84 |

IQR: interquartile range, SD: standard deviation, COPD: chronic obstructive pulmonary disease, LMWH: low molecular weight heparin. <sup>1</sup>Systemic autoimmune disease, 4 patients; Multiple Sclerosis 2, patients; lymphoproliferative disease 2 patients; solid organ transplantation, 2 patients; primary immunodeficiency, myasthenia gravis and inflammatory bowel disease, 1 patient each. <sup>2</sup>Upon discharge, 15 cases and 14 controls received LMWH in prophylactic doses

Neither glucocorticoids nor anticoagulants prescribed at hospital discharge were associated with a lower readmission rate. (Table 2). Seventeen cases and 16 controls received anticoagulant treatment. Anticoagulant treatment consisted of low molecular weight heparin (LMWH) at prophylactic doses in 88.2% of cases and 87.5% of controls, respectively. No difference in analytical data at admission or discharge was evident between the two groups (Table 3). Neutrophil/lymphocyte ratio at hospital admission tended to be higher in cases than in controls (p=0.06). Twenty-nine readmitted patients had a new PCR assay at re-entry. Viral RNA was detected in 15 of them (51.7%).

**Table 3.** First and last laboratory analysis results of patients with COVID-19.

|  | Cases | Controls | p |
| --- | --- | --- | --- |
| At hospital admission (median, IQR) |  |  |  |
| C Reactive Protein, mg/L | 34.5 (15.8-100.2) | 37.8 (8.7-70.0) | 0.28 |
| D-Dimer, ng/ml | 1.08 (0.48-1.94) | 0.65 (0.36-1.54) | 0.09 |
| Lactate Dehydrogenase, units/L | 266.5 (216-322) | 251.0 (213-323) | 0.62 |
| Ferritin, ng/mL | 756 (302-2081) | 603 (370-869) | 0.55 |
| Lymphocyte count cel/uL | 970 (670-1460) | 1090 (870-1430) | 0.31 |
| Neutrophil count cel/uL | 4390 (3070-7300) | 3960 (2770-6110) | 0.30 |
| Neutrophil-to-lymphocyte ratio | 4.32 (2.82-7.40) | 3.23 (2.30-5.31) | 0.06 |
| Pre-discharge (median, IQR) |  |  |  |
| C Reactive Protein, mg/L | 15.7 (5.2-49.5) | 15.3 (4.0-35.2) | 0.45 |
| D-Dimer, ng/ml | 0.89 (9.38-1.44) | 0.60 (0.39-1.37) | 0.41 |
| Lactate Dehydrogenase, units/L | 225 (178-278) | 241 (192-286) | 0.50 |
| Ferritin, ng/mL | 670 (416-1434) | 469 (308-876) | 0.11 |
| Lymphocyte count cel/uL | 1070 (600-1700) | 1360 (900-1620) | 0.09 |
| Neutrophil count cel /uL | 3820 (2100-5700) | 3940 (2865-5480) | 0.53 |
| Neutrophil-to-lymphocyte ratio | 3.13 (1.90-8.43) | 3.01(1.74-5.68) | 0.35 |

IQR: interquartile range.

### Description of the indication for admission at initial admission and readmission

Tables 4 shows the reason for the first admission in cases and controls, and table 5 compares the indication of admission between the first and second hospitalization for cases. Although in cases and controls pneumonia was the most frequent cause of admission, it was more common among controls (p=0.05, Table 4).

**Table 4.** Cause of hospital admission in cases (first admission) and controls.

| Condition | Cases<br>n=61 | Controls<br>n=61 | p |
| --- | --- | --- | --- |
| Pneumonia | 53 (86.9) | 59 (96.7) | 0.05 |
| Heart failure | 2 (3.3) | 1 (1.64) | 0.55 |
| Bacterial superinfection | 5 (8.2) | 4 (6.7)<sup>1</sup> | 0.75 |
| Acute kidney failure | 1 (1.6) | 0 (0.0) | 0.32 |

1. In three patients there was lung co-infection

**Table 5.** Cause of hospital admission and readmission in cases.

| Condition | initial admission<br>(n=61) | Readmission (n=61) |
| --- | --- | --- |
| Pneumonia | 53 (86.9) | 34 (55.7)<sup>1</sup> |
| Pulmonary thromboembolism |  | 8 (13.1)<sup>1</sup> |
| Heart failure | 2 (3.3) | 6 (9.8) |
| Bacterial infection | 5 (8.2) | 4 (6.6) |
| Acute kidney failure | 1 (1.6) | 2 (3.3) |
| Deep vein thrombosis |  | 1 (1.6) |
| Lower limb arterial thrombosis |  | 1 (1.6) |
| Myocardial acute infarction |  | 1 (1.6) |
| Severe bleeding |  | 1 (1.6) |
| Diabetes, hyperglycaemic hyperosmolar<br>state |  | 1 (1.6) |
| Generalized oedema |  | 1 (1.6) |
| Threatened miscarriage |  | 1 (1.6) |

<sup>1</sup> p<0.001.

All cases that presented pulmonary thromboembolism or deep vein thrombosis were diagnosed during the second admission (p=0.001). The motive for readmission in 10 patients (16.4%) was a thrombotic event in venous or arterial territory. Among them, 7 patients had no anticoagulant medication recommended at discharge (p=0.07).

## Discussion

The readmission rate shown in this study was low, suggesting that discharge decisions were frequently appropriate and the prognosis for most patients was favourable. Immunocompromised patients and those who presented fever within 48 hours prior to discharge were at increased risk of readmission.

These results are similar to those of other published studies, with a readmission rate during the first weeks between 2 and 4% (6–7). The vast majority of COVID-19 patients who are discharged are not readmitted, suggesting that they are not discharged hastily. In addition, the readmission rate of COVID-19 patients appears to be lower than that observed in conventional patients treated in internal medicine wards. This fact may be related to the more advance age and relevant comorbidity in the latter (9). In any case, further research would be desirable to analyse how to shorten the hospital stay even further without significantly increasing readmissions.

### Clinical characteristics of the patients who are readmitted

Our immunocompromised patients were at increased risk for re-admission. The effect of immunosuppression on the clinical course of SARS-Cov-2 infection is not well known. Long-term use of glucocorticoids can cause atypical clinical presentation with a longer incubation period, which could reduce the clinical suspicion of this infection (10). Likewise, mild cases have been described in relation to the mitigation of cytokine storm by the previous use of immunosuppressive drugs (11). In fact, some immunocompromised patients are receiving treatment with interleukin inhibitors such as tocilizumab or anakinra, which are drugs that have been successfully used in patients with COVID-19. On the other hand, it has been observed that transplanted patients have a clearly worse prognosis than other types of patients (12). These discordant findings may be due to the different effects of each type of immunosuppression on innate and acquired immunity and the uncertainty of the role of viral replication in the prognosis of the disease (12–14). Until the prognosis and evolution of immunocompromised patients with COVID-19 is better known, it is advisable to carefully decide the moment of hospital discharge in these patients.

Patients with hypertension in our series showed a trend towards a more frequent readmission (p=0.07). Other studies have reported a higher risk of readmission in patients with hypertension, which could be related to the more severe disease of COVID-19 in hypertensive patients (1,7). Our study did not show an increased risk of readmission in COPD patients, which, according to Sulaiman et al., (7) seems to be expected from the hypoxemia associated with respiratory infections in these patients. Patients who present with fever during the 48 hours prior to discharge are at increased risk of readmission and should be followed closely. Differences in age between cases and controls were not analysed, due to age-adjusted case-control matching, but previous study showed no difference in age in terms of risk of readmission (7)

Of note, analytical alterations (LDH, C-reactive protein, neutrophil/lymphocyte ratio, and D-dimer) were not useful in predicting readmission. There was a trend towards higher D-dimer and neutrophil/lymphocyte ratios on admission and lower lymphocyte counts on discharge in patients who were readmitted (Table 3). This result has also been reported by other authors and reinforces the belief that clinical variables (such as fever or respiratory failure) are more important than analytic ones (such as the degree of lymphopenia or the plasma level of C-reactive protein) for decision making in this disease. (7).

### Description of the cause for admission at initial admission and readmission

The immobilization due to asthenia and malaise, hypoxia, coagulopathy and endothelial viral damage that characterize COVID-19 predispose patients to venous thromboembolic disease (VTE) (15,16). Nevertheless, cases of VTE usually appear progressively during the 4 weeks after the first arrival in hospital, which may explain why it is a more frequent cause of admission in the second than in the first admission (16) The use of prophylactic LMWH in our institution increased as we became more aware of the risk of VTE. Patients who were readmitted for vascular thrombotic problems tended to be discharged from the first admission without anticoagulant drugs (p=0.07). However, the prophylactic strategy with LMWH has not been successful in all cases (17). There is still a need to better define the risk estimate and the dose and duration of LMWH prophylaxis in COVID-19 (15–17).

Moreover, treatment of patients during initial admission with intravenous fluids, glucocorticoids and anticoagulants may cause worsening of chronic diseases such as heart failure or diabetes and end up causing readmissions. The careful use of medication and optimal medical care is a clear objective in the management of these patients.

This study has several limitations that should be highlighted. First, the small sample size may have prevented the identification of differences in some variables. Secondly, it is a single-centre study having its own diagnostic and therapeutic peculiarities. Thirdly, we cannot completely exclude that some of the control patients had been admitted to a private hospital, whose information is not collected in the public health informatics system. However, this is not likely to have occurred. And finally, it should be noted that the demand for hospital admissions and the learning curve of the disease has changed over the course of the epidemic, which may have had a varying impact on patient discharge.

In summary, we report a low rate of readmission after discharge from hospital for COVID-19. Immunocompromised patients and those presenting with fever during the 48 hours prior to discharge are at greater risk of readmission to hospital. Given the possibility of further outbreaks of the disease, further research should be encouraged to refine the risk factors for hospital readmission that could help to safely discharge these patients.

## Data Availability

All data concerning the research are available by contacting the corresponding author

## Ethical statement

This study was approval the local Clinical Research Ethics Committee (CEIC). All patients gave their consent to participate in the study.

## Conflicts of interest

The authors declare that they do not have any conflict of interest related to the content of the article.

## Funding

This study did not receive any funding.

## Notes

### Competing Interest Statement

The authors have declared no competing interest.

### Clinical Trial

NA

### Funding Statement

None

## References

1. Cao H, Ruan L, Liu j, Liao W. The Clinical Characteristic of Eight Patients of COVID-19 With Positive RT-PCR Test After Discharge. J Med Virol 2020. Online ahead of print.

2. Dafer RM, Osteraas ND, Biller J. Acute Stroke Care in the Coronavirus Disease 2019 Pandemic. J Stroke Cerebrovasc Dis 2020 Online ahead of print.

3. Al-Shamsi HO, Alhazzani W, Alhuraiji A, Coomes EA, Chemaly RF, Almuhanna M, et al. A Practical Approach to the Management of Cancer Patients During the Novel Coronavirus Disease 2019 (COVID-19) Pandemic: An International Collaborative Group. Oncologist. 2020 Online ahead of print.

4. Yu L. Standards and Follow-up Plan for COVID-19 Patients. In Handbook of COVID-19 Prevention and Treatment, Yu L editor. 2020: 45–47.

5. Richardson S, Hirsch JS, Narasimhan M, et al. Presenting Characteristics, Comorbidities, and Outcomes Among 5700 Patients Hospitalized With COVID-19 in the New York City Area. JAMA. 2020 Apr. DOI: 10.1001/jama.2020.6775.

6. Wang X, Xu H, Jiang H, Wang L, Lu C, Wei X, et al. The Clinical Features and Outcomes of Discharged Coronavirus Disease 2019 Patients : A Prospective Cohort Study. QJM 2020 Online ahead of print.

7. Sulaiman S, Richter F, Fuster V, De Freitas J, Naik N, Sigel K, et al. Characterization of Patients Who Return to Hospital Following Discharge from Hospitalization For COVID-19. MedRxiv preprint doi: https://doi.org/10.1101/2020.05.17.20104604

8. Real-Time RT-PCR Panel for Detection 2019-Novel Coronavirus Centers for Disease Control and Prevention, Respiratory Viruses Branch, Division of Viral Diseases. Accessible in https://www.who.int/docs/default-source/coronaviruse/uscdcrt-pcr-panel-for-detection-instructions.pdf?sfvrsn=3aa07934_2

9. Zapatero A, Barba R, Marco J, Hinojosa J, Plaza S, Losa JE, et al. Predictive model of readmission to internal medicine wards. Eur J Intern Med 2012; 23: 451–6

10. Han Y, Jiang M, Xia D, He L, Lv X, Liao X, et al. COVID-19 in a patient with long-term use of glucocorticoids: A study of a familial cluster. Clin Immunol. 2020 214: 108413.

11. Spezzani V, Piunno A, Iselin HU. Benign COVID-19 in an immunocompromised cancer patient – the case of a married couple. SwissMedWkly 2020; 150: w20246

12. Fernández-Ruiz M, Andrés A, Loinaz C, Delgado JF, López-Medrano F, San Juan R, et al. COVID-19 in solid organ transplant recipients: A single-center case series from Spain. Am J Transplant 2020 Online ahead of print.

13. Huang C, Wang Y, Li X, Ren L, Zhao J, Hu Y, et al. Clinical features of patients infected with 2019 novel coronavirus in Wuhan, China. Lancet 2020; 395 (10223): 497–5

14. Cavagna L, Bruno R, Zanframundo G, Gregorini M, Seminari E, et al. Clinical presentation and evolution of COVID-19 in immunosuppressed patients. Preliminary evaluation in a North Italian cohort on calcineurin-inhibitors based therapy. MedRxiv. doi: https://doi.org/10.1101/2020.04.26.20080663

15. Tang N, Bai H, Chen X, Gong J, Li D, Sun Z. Anticoagulant treatment is associated with decreased mortality in severe coronavirus disease 2019 patients with coagulopathy. J Thromb Haemost 2020; 18:1094–9.

16. Klok FA, Kruip MJHA, van der Meer NJM, Arbous MS, Gommers D, Kant KM, et al. Confirmation of the high cumulative incidence of thrombotic complications in critically ill ICU patients with COVID-19: An updated analysis. Thromb Res 2020 Online ahead of print.

17. Poggiali E, Bastoni D, Ioannilli E, Vercelli A, Magnacavallo A. Deep Vein Thrombosis and Pulmonary Embolism: Two Complications of COVID-19 Pneumonia? Eur J Case Rep Intern Med 2020; 7:001646

